# The association between age, COVID-19 symptoms, and social distancing behavior in the United States

**DOI:** 10.1101/2020.04.19.20065219

**Authors:** David Canning, Mahesh Karra, Rashmi Dayalu, Muqi Guo, David E. Bloom

## Abstract

**Background:** Public health authorities recommend that people practice social distancing, especially if they have symptoms of coronavirus disease (COVID-19), or are older and more at risk of serious illness if they become infected. We test the hypothesis that these groups are following these recommendations and are more likely to undertake social distancing.

**Methods:** We conducted an open online survey of 4,676 U.S. adults aged 18 and older between April 4 and April 7, 2020. We model the effects of age and common COVID-19 symptoms in the last two weeks on going out of the home for non-healthcare reasons the day before taking the survey, using a logistic model and the number of close contacts (within 6 feet) that respondents had with non-household members, using a Poisson count model. Our models control for several covariates, including other flu-like symptoms, sex, education, income, whether the respondent worked in February, household size, population density in the respondent’s ZIP code, state fixed effects, and the day of completion of the survey. We also weight our analyses to make the sample representative of the U.S. adult population.

**Findings:** About 52 percent of the adult United States population went out of their home the previous day. On average, adults had close contact with 1.9 non-household members. We find that having at least one COVID-19 symptom (fever, dry cough, or shortness of breath) *increased* the likelihood of going out the previous day and having additional close contacts with non-household members; however, the estimates were not statistically significant. When disaggregating our analysis by COVID-19 symptoms, we find no strong evidence of greater social distancing by people with a fever or cough in the last two weeks, but we do find that those who experienced shortness of breath have fewer close contacts, with an incidence rate ratio (IRR) of 0.49 (95% CI: 0.30–0.78). Having other flu-like symptoms reduces the odds of going out by 0.32 (95% CI: 0.18–0.60) and the incidence rate of having close contacts by 42 percent (IRR = 0.58; 95% CI: 0.38–0.88). We find that older people are just as likely to leave their homes as younger people, but people over the age of 50 had less than half the predicted number of close contacts than those who were younger than 30. Our approach has the limitation that the survey sample is self-selected. Our findings may therefore be subject to selection bias that is not adequately controlled for by weighting. In addition, the possibility exists of confounding of the results due to omitted variable bias.

**Conclusions:** We provide evidence that older people are having significantly fewer close contacts than younger people, which is in line with the public health authorities’ recommendations. We also find that people experiencing shortness of breath are practicing more intense social distancing. However, we find that those with two other common COVID-19 symptoms, fever and dry cough, are not engaging in greater social distancing, suggesting that increased targeting on relevant symptoms, and messaging, may be required.

## Introduction

The rapid spread of coronavirus disease 2019 (COVID-19) and the fear of massive numbers of fatalities has led to a worldwide call for social distancing measures to slow the spread of the epidemic [1, 2]. While it is unclear how long these measures will have to stay in place [3], recommendations or requirements to socially distance have now been implemented in many countries [4, 5], including the United States [6]. In addition to federal-level recommendations from the Centers for Disease Control and Prevention (CDC), many states have implemented stricter regulations that limit which businesses can operate and restrict the ability of people to leave their homes and to meet in groups.

Information on compliance with these recommendations and regulations is required to understand if they are being followed and whether they are likely to work. This information is also useful for epidemiological and economic models that use estimates of social distancing compliance and effectiveness to assess the likely course of the epidemic and economic costs under different policy scenarios [7-10].

Several studies have investigated compliance with social distancing regulations and recommendations due to COVID-19. Based on location tracking data from cell phones, political party affiliation at the county level in the United States has been found to be predictive of movement [11, 12]. Survey data have found that compliance increased with empathy for vulnerable groups in the United States, United Kingdom, and Germany [13], while reported willingness to comply was higher for shorter rather than longer expected periods of restriction in Italy [14]. However, a recent international open web-based survey of 324 individuals found that fear of contracting the virus was the only strong predictor of social distancing behavior [15].

We focus on whether or not individuals with common symptoms of COVID-19, and older people who are more vulnerable to illness, are practicing increased social distancing, as is recommended. We leave the issue of why individuals may or may not comply with the recommendation to future research; empathy for others and fear of infection are important mechanisms that may be mediating our results.

We investigate the effect of having a common symptom of COVID-19 on social distancing behavior. The World Health Organization (WHO) identifies fever, tiredness, and dry cough as the most common symptoms of COVID-19 infection [16]. The WHO lists other symptoms, including shortness of breath, aches and pains, and sore throat, with few people reporting diarrhea, nausea, or a runny nose. The WHO advises that people with fever, cough, and difficulty breathing should seek medical attention. The CDC identifies fever, cough, and shortness of breath as common symptoms of COVID-19 infection [17]. The CDC recommends that everyone should stay at home as much as possible and avoid close contact with others (within 6 feet) to prevent infection. The CDC also recommends that people who are sick should stay home except to get medical care [6] and that those with higher risk of severe illness, such as the elderly, should take extra precautions [18]. We use a combination of these criteria to derive our measures of common symptoms of COVID-19 and recommended social distancing behavior in the United States.

Some evidence indicates that other symptoms, in particular the loss of taste and smell, have a high specificity for COVID-19 [19]. However, these symptoms are not yet included in the public list of common symptoms that public health agencies are reporting, or are using as a basis for social distancing recommendations.

We undertook an open-access, web-based survey that was hosted on two sites: one at the Program on the Global Demography of Aging (PGDA) at Harvard University and one at the Global Development Policy Center (GDP Center) at Boston University. Both web portals led viewers to the same survey. Both universities advertised the survey through various channels, including social media (Facebook, Twitter, Instagram, and LinkedIn), e-mail announcements and newsletters, and website advertisements. For this study, we use data collected from respondents over a 4-day period, from April 4 to April 7, 2020. The survey was designed to take less than 5 minutes to encourage participation and completion [20]. Our survey was open access to elicit a large number of responses in a short period of time, which would allow us to obtain rapid information on social distancing behaviors. Online surveys have been found to offer higher-quality responses than telephone surveys [21].

We model the effects of age and common COVID-19 symptoms in the last two weeks on respondents’ going out of the home, for reasons other than for healthcare, on the day before they took the survey. We model this binary outcome using a multivariate logistic regression. We also model the effect of age and common COVID-19 symptoms in the last two weeks on the number of close contacts (within 6 feet) that respondents had on the day before they took the survey. We model this count variable using a Poisson regression. In both regressions, we control for factors that might affect a respondent’s social distancing behavior, such as other flu-like symptoms, sex, education, income, whether the respondent worked in February, household size, population density in the respondent’s ZIP code, state fixed effects, and the day of completion of the survey.

Relative to the general U.S. population, our sample is heavily skewed toward people with higher education levels, younger adults, and women. We use weights that are derived from the 2018 American Community Survey [22] to make our sample representative of the United States population. While the need to use weights to make descriptive statistics representative of the population is clear, using weights in regression models may not always be desirable [23]. We present weighted regression results for our statistical analyses in this paper, but also present the same regression analyses without weights in an online appendix. The conclusions based on these unweighted regressions are similar to those we find when using the weights. While weights can make the sample representative of the population, evidence indicates that nonprobability-based web surveys are less representative than probability-based surveys even after weighting [21].

## Methods

### Data Collection

We collected data via an online Qualtrics-based survey with portals on the Harvard PGDA website (https://www.hsph.harvard.edu/pgda/covid/) and the Boston University GDP Center website (http://www.bu.edu/gdp/research/human-capital-initiative/covid-19-symptoms-social-distancing-web-survey/). The survey was designed to be short to encourage completion and rapid data collection; the median time respondents spent to complete the survey was 3.6 minutes. The survey questionnaire collected basic demographic information, recent work experience, symptoms of COVID-19 and some other health conditions, and social distancing behavior. A copy of the questionnaire is available in our online appendix.

### Ethical Considerations

The survey was open only to residents of the Unite States over age 18. All subjects gave informed consent after reading a conscript script. None of the data collected included individual identifiers. The questionnaire contained a link to a CDC website for advice on what to do if people had symptoms of a COVID-19 infection, and another CDC web site, and the number of a 24 hour telephone hot-line, with advice for anyone feeling stress or anxiety. The study received a determination of exemption from human subjects research approval from the institutional review boards at Harvard University (protocol number IRB20-0548) and Boston University (protocol number 5551X).

### Outcomes

Our study has two outcome measures. The first is based on the question: “Did you leave your home yesterday?” with answers yes (coded 1) or no (coded 0). The CDC recommends people to not leave their home if they are sick, with the exception of seeking healthcare. For people who did leave their home, we also ask: “What were your reasons for going out yesterday?” This question is followed by a list of reasons (see questionnaire in online appendix), one of which is “healthcare/visit to doctor or pharmacy.” We code those people who left their home exclusively for healthcare as zero because they were following the CDC recommendations for social distancing. Our second outcome measure is based on the question: “Excluding members of your household, how many people in total did you come into close contact with (within 6 feet) yesterday?” This question aims to capture those contacts that a respondent had as a result of going outside the home and those contacts who came into the respondent’s home in the last day. The number of contacts 0,1,2,3, and 4 are reported as integers, and we record ranges for larger numbers (see Table 1). To calculate the number of contacts, we use the mid-point of the range and top code responses to 30 contacts.

**Table 1.** Descriptive characteristics of respondents.

| Characteristic | Total Population<br>(N=4,676) |
| --- | --- |
| **Categorical variables** | **n (%)** |
| Any COVID-19 symptoms in last two weeks<sup>1</sup> | 953 (20.4%) |
| Fever | 172 (3.7%) |
| Dry cough | 698 (14.9%) |
| Shortness of breath | 349 (7.5%) |
| Other flu-like symptoms | 298 (6.4%) |
| Went out yesterday<sup>2</sup> | 2,587 (55.3%) |
| Number of close contacts yesterday<sup>3</sup> |  |
| 0 | 2,877 (61.5%) |
| 1 | 564 (12.1%) |
| 2 | 384 (8.2%) |
| 3 | 222 (4.7%) |
| 4 | 178 (3.8%) |
| 5 - 10 | 279 (6.0%) |
| 11 - 20 | 99 (2.1%) |
| More than 20 | 73 (1.6%) |
| Sex |  |
| Female | 3,712 (79.4%) |
| Male | 964 (20.6%) |
| Age groups |  |
| 18-29 | 738 (15.8%) |
| 30-39 | 1,294 (27.7%) |
| 40-49 | 1,144 (24.5%) |
| 50-59 | 784 (16.8%) |
| 60-69 | 449 (9.6%) |
| ≥70 | 267 (5.7%) |
| Education level |  |
| Less than high school | 3 (0.1%) |
| High school | 89 (1.9%) |
| Some college | 321 (6.9%) |
| 2-year associate degree from a college or university | 171 (3.7%) |
| 4-year college or university degree | 1,134 (24.3%) |
| Some postgraduate or professional schooling, no postgraduate degree | 450 (9.6%) |
| Postgraduate or professional degree | 2,508 (53.6%) |
| Individual income in 2019 |  |
| Less than \$20,000 | 533 (11.4%) |
| \$20,000 to less than \$50,000 | 850 (18.2%) |
| \$50,000 to less than \$100,000 | 1,495 (32.0%) |
| \$100,000 or more | 1,798 (38.5%) |
| Have paid work in February 2020 | 3,811 (81.5%) |
| Household size (including the respondent) |  |
| 1 | 531 (11.4%) |
| 2 | 1,452 (31.1%) |
| 3 | 966 (20.7%) |
| 4 | 1,135 (24.3%) |
| ≥5 | 592 (12.7%) |
| Day of survey completion |  |
| Saturday April 4, 2020 | 112 (2.4%) |
| Sunday April 5, 2020 | 2,148 (45.9%) |
| Monday April 6, 2020 | 1,945 (41.6%) |
| Tuesday April 7, 2020 | 471 (10.1%) |
| **Continuous variables** | **mean (standard deviation)** |
| Natural logarithm of population density per square mile | 7.8 (1.6) |
| Weight of U.S. population by sex, age, and education | 47,563.8 (18,9871.8) |

Notes: <sup>1</sup>Any COVID-19 symptom includes fever, dry cough, or shortness of breath. All symptoms are measured over the last two weeks; <sup>2</sup>Yesterday refers to the day before completing the survey; Going out does not include those who went out only for health care; <sup>3</sup> Close contact means coming within 6 feet of a non-household member.

### Primary Exposures

Our primary exposure measures are age and common symptoms of COVID-19. Age is reported in the survey in single years from age 18 and up; only respondents who are 18 and older are eligible to participate in the survey. For our analysis, we divide age into six groups: 18–29, 30–39, 40–49, 50–59, 60–69, and 70 and older. For common symptoms of COVID-19, we ask respondents: “Have you experienced any of the following symptoms in the past two weeks?” The common symptoms of COVID-19 that we focus on are fever, dry cough, and shortness of breath. We also ask respondents about “other flu-like symptoms” that they might have experienced in the past two weeks. For each symptom, respondents can either answer “yes,” “no,” or “don’t know.” We code “no” and “don’t know” responses from respondents as 0 and “yes” responses as 1. Our results are robust to dropping “don’t know” responses from the analysis. We create a dummy variable for having at least one common COVID-19 symptom (i.e., fever, dry cough, or shortness of breath) in the last two weeks. We also analyze the effect of each symptom separately.

### Statistical Analyses

Leaving the home yesterday (other than for healthcare) is a binary variable, and we analyze this outcome using a logistic regression model. We report coefficient estimates from the logistic regression as odds ratios. The number of close contacts yesterday is a count variable, and we analyze this outcome using a Poisson regression model. We report coefficient estimates from the Poisson regression as incidence rate ratios. For both logistic and Poisson regression models, we use heteroskedastic robust standard errors to calculate confidence intervals.

We control for various individual- and household-level covariates in our regression analysis. As well as common COVID-19 symptoms, we control for “other flu-like symptoms.” We also control for respondent sex (male versus female), reported individual income in 2019 (less than $20,000, $20,000–$49,999, $50,000–$99,999, and more than $100,000), and current household size (in integers for 1,2,3,and 4 and then the group 5 or more). We include level of education, which is reported in seven categories in our survey; for our analysis, we collapse this variable into two categories (some college or more versus high school or less).

In addition to these individual- and household-level variables, we also control for some community-level variables. The first is the log (1+ population density per square mile) in the respondent’s ZIP code. Population density may affect both the perceived safety of leaving the home and influence the number of non-household members that a respondent may come into contact with. Second, we add 52 fixed effects for the 50 states plus the District of Columbia and Puerto Rico, which adjust the results for the variation in social distancing policies by state. In addition, our survey was implemented over four days, and we add day-of-survey fixed effects, which adjust for variation in behavior by the day of the week.

We exclude several variables that may be related to social distancing but that are also potential mediators of the effect from our models. For example, one reason for going out in the last two weeks is for work, but we do not control for this variable because not working outside the home may be a consequence of social distancing rather than a cause. We do, however, control for whether the respondent worked for pay in February 2020 because this variable may be a driver of continued work outside the home.

We weight the survey using data from the 2018 American Community Survey (ACS) to make our sample representative of the U.S. population. Using ACS survey data, we calculate the population by sex, age, and education groups. We divide the population into sex (male and female), age groups (18–29, 30–39, 40–49, 50 –59, 60–69, and 70+), and two education groups (some college or more and high school or less). This division gives us 2 × 6 × 2 = 24 groups. We use the ratio of these population numbers to the number in the corresponding sex, age, and education group in our survey as weights. Both our descriptive statistics and our regression results use weights. In our online appendix, we present all our results without the use of weights. Arguments exist both for and against weighting the sample [23], and some readers may prefer the unweighted results.

We use our logistic regression results to predict the probability that a person left the home yesterday by age group and COVID-19 symptoms. These predictions are made at the means of the other variables. For example, the predicted probability of leaving the home yesterday for someone without COVID-19 symptoms is based on their having the average value of all the other covariates in the model. Similarly, we use the Poisson model to calculate the expected number of close contacts yesterday for different age groups and symptoms, fixing other covariates at their mean values.

## Results

Table 1 presents unweighted descriptive statistics for our sample. We present data from 4,676 responses collected over 4 days. Slightly more than 20 percent of our sample reports at least one common symptom of COVID-19 in the last two weeks. By far the most common of these reported symptoms is a dry cough (14.9 percent), while the proportions of respondents who report a fever (3.7 percent) and shortness of breath (7.5 percent) are lower. Other flu-like symptoms were reported by 6.4 percent of the sample. About 55 percent of respondents in the sample report having left their home yesterday (the day before they completed the survey). Only 49 percent of respondents report any close contact with any non-household member in the previous day. However, 3.7 percent of respondents report having had more than 10 close contacts in the previous day.

Table 1 shows that our sample skews heavily toward highly educated groups, is predominantly female, and comprises mainly younger adults. Slightly more than 70 percent of the sample report making more than $50,000 a year, while 81 percent report working for pay in February. The most common household size is 2, with around 13 percent of the sample being top coded as having 5 or more household members.

Figure 1 shows the estimated percentages of people with different symptoms in the last two weeks, with the percentages being weighted to make the sample representative of the U.S. adult population. The error bars in the figure indicate the 95 percent confidence intervals. The presented rates are close to those that are reported in the raw data for the sample in Table 1. Slightly less than 20 percent of adults have any common COVID-19 symptom, and the most common of the three symptoms, by far, is a dry cough.

**Figure 1.**
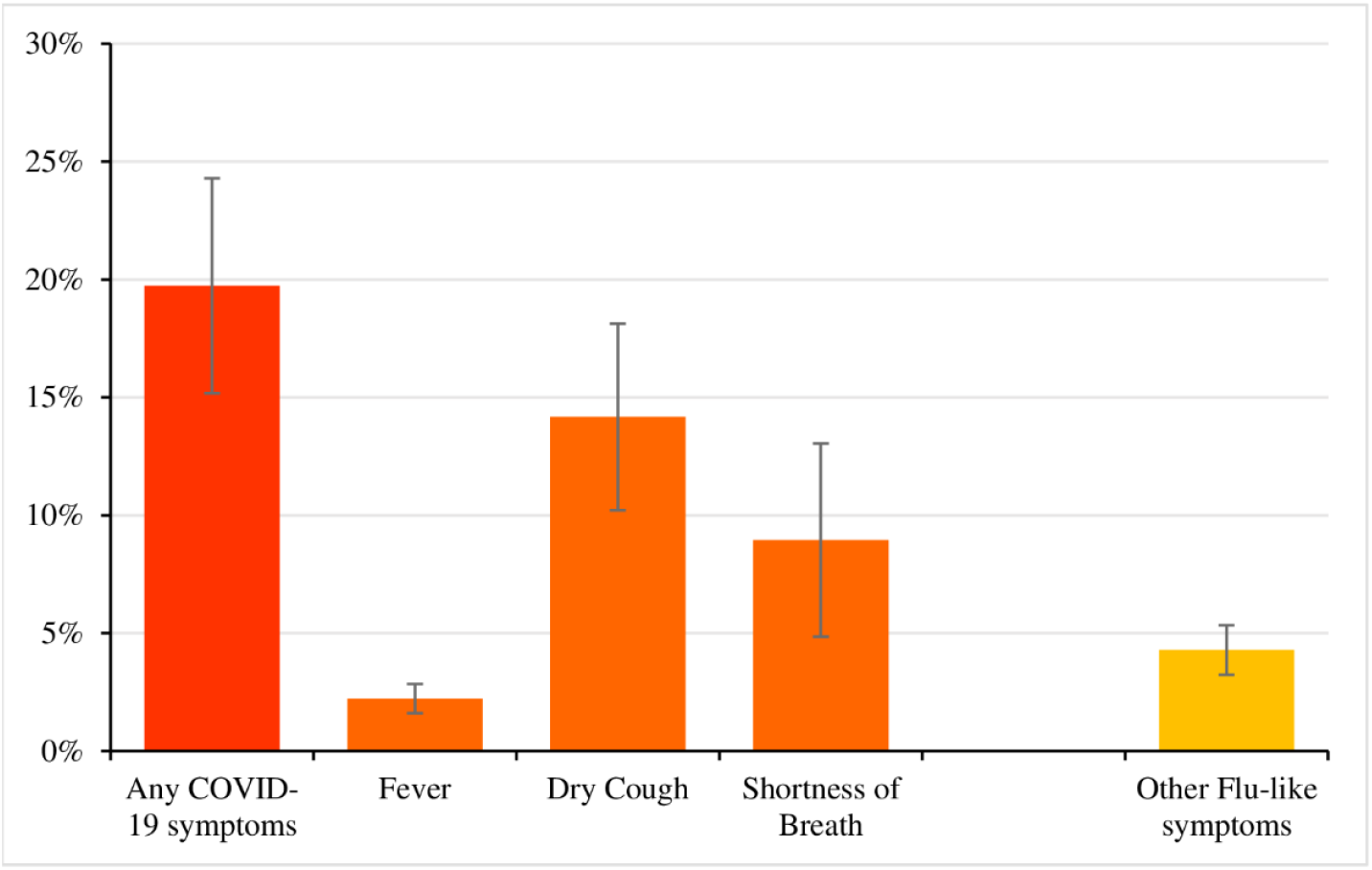
Prevalence of COVID-19 symptoms in last two weeks. Notes: (A) Any COVID-19 symptoms include fever, dry cough, or shortness of breath; (B) Error bars represent 95 percent confidence intervals of symptom prevalence rate; (C) Values were weighted to be representative of U.S. population by age, sex, and education.

In Table 2, Model 1 reports estimates for the determinants of going out of the home the day before the survey. We use sample weights, the coefficient estimates are reported as odds ratios, and the confidence intervals are calculated using heteroskedastic robust standard errors. We estimate the effect of any common symptom of COVID-19 in the last two weeks on the likelihood of leaving the home. Surprisingly, we find a slightly higher likelihood of going out for those respondents who report having at least one symptom relative to those respondents without any symptoms. We find an odds ratio of 1.38, but the 95% confidence interval (95% CI) is (0.89–2.14), which spans the null of an odds ratio of 1, and the estimate is not statistically significant at the 5 percent level. However, having other flu-like symptoms is associated with a significant decrease in the likelihood of going out, with an estimated odds ratio of 0.34 (95% CI: 0.20–0.59). None of the age group coefficients are statistically significant, indicating a lack of any strong trend in the likelihood of going out by age. We also find no significant effects of education, or income level. However, working for pay in February 2020 is predictive of going out, with an odds ratio of 1.89 (95% CI: 1.19–3.00). Household size is not associated with the probability of going out, but people who live in ZIP codes with a higher population density are more likely to go out than those living in areas with a lower population density.

**Table 2.**
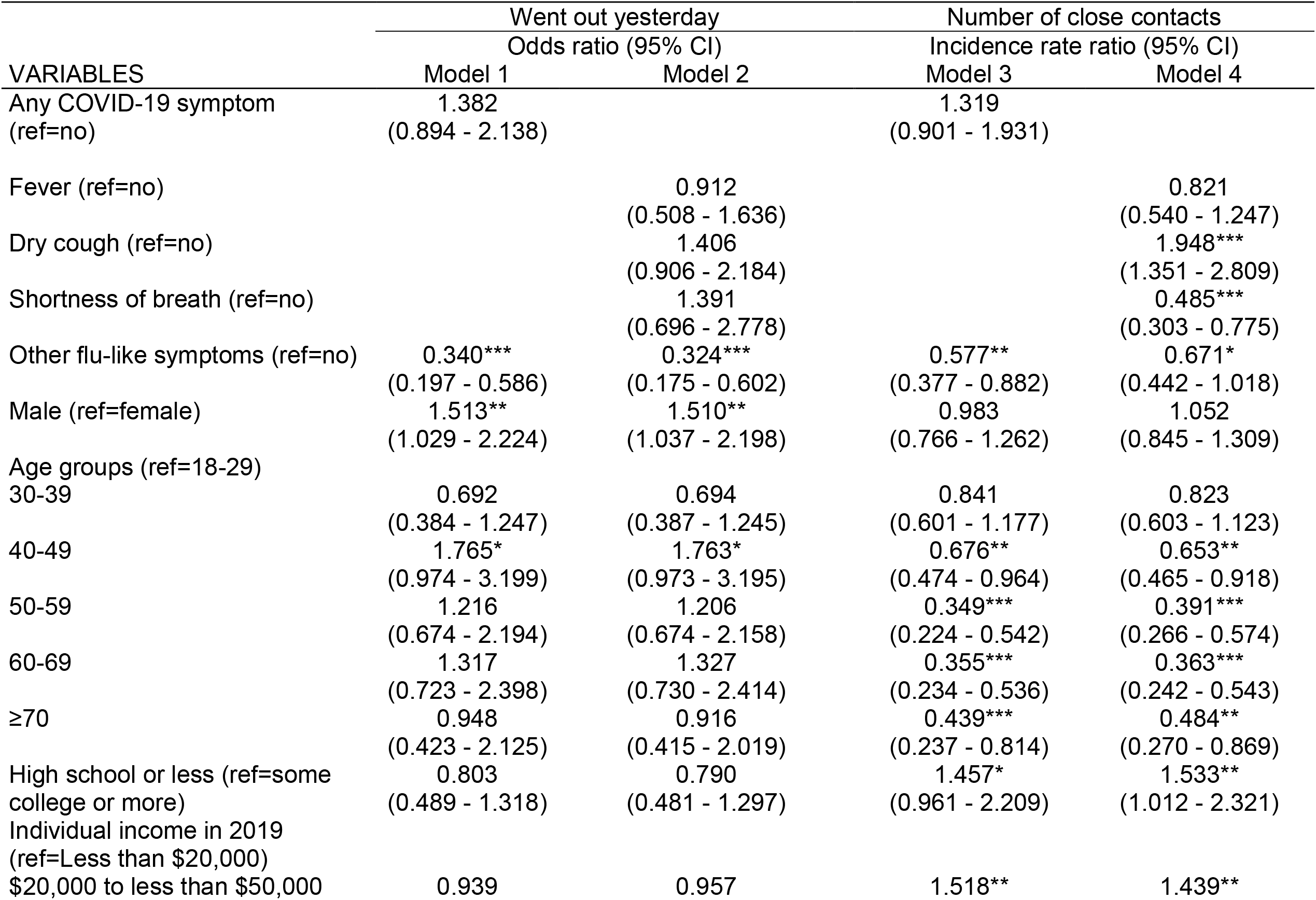
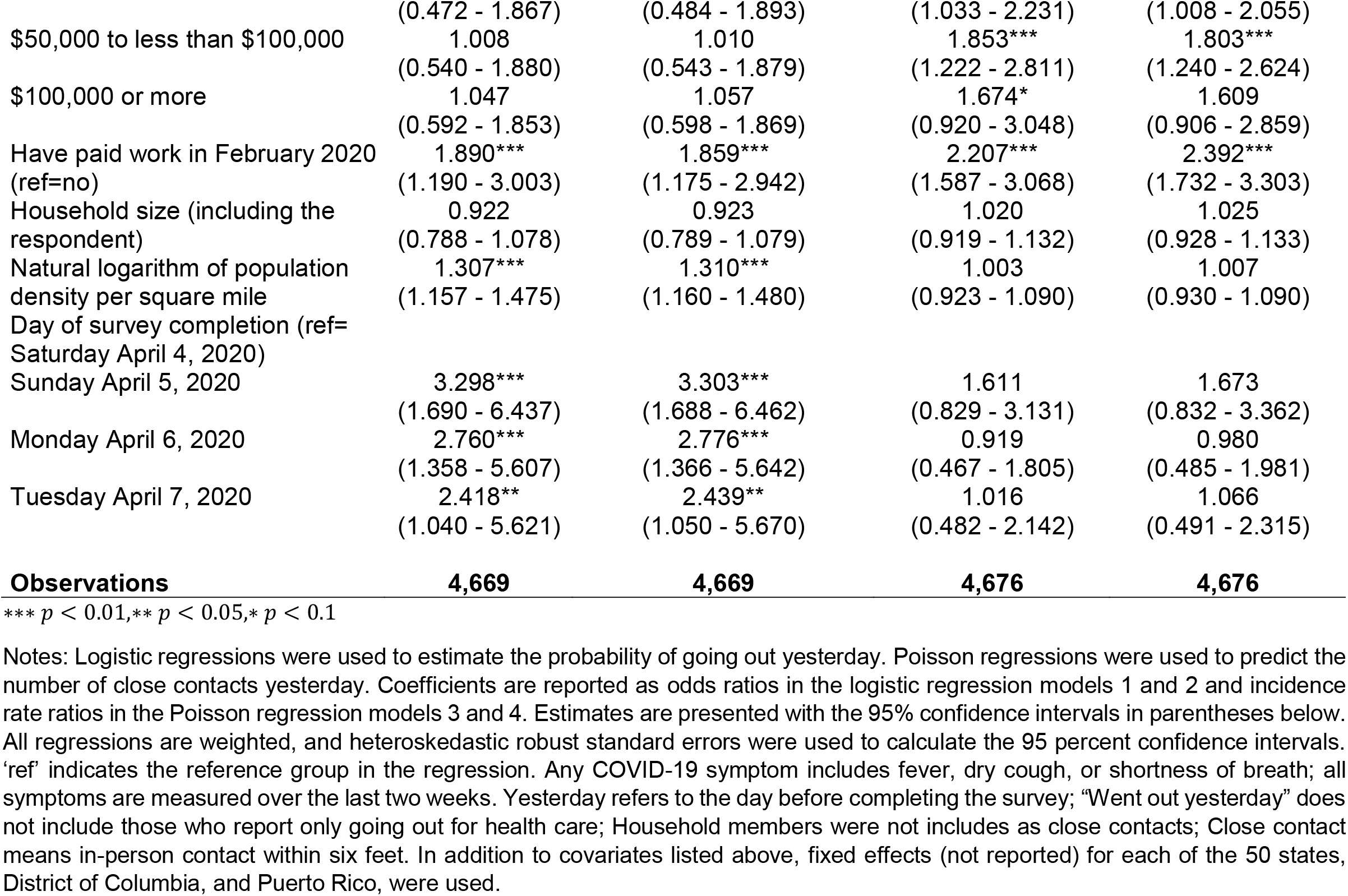
**Multivariate analysis of going out, and number close contacts, the day before the survey**

In Model 2, we estimate the same regression as in Model 1, but separate out the effect of the three symptoms, fever, dry cough, or shortness of breath. We find no statistically significant effects of the three symptoms taken separately in Model 2. The estimates of the effect of the other factors, including the large reduction in the odds of going out for people with other flu-like symptoms, remain very similar to the findings in Model 1.

We can use Models 1 and 2 to predict the probability that a person with particular symptoms goes out. For example, we use Model 1 to find the predicted probability that someone with no symptoms goes out, given that their other characteristics are at the mean value for the population. This predicted probability is shown in the top bar in Figure 2, along with a 95% confidence interval for the predicted value. We can compare this probability with the predicted probability that someone with at least one common COVID-19 symptom goes out, again calculated at the mean value of the other variables, shown in the second bar from the top in Figure 2. The predicted probability for someone with a COVID-19 symptom is slightly higher than for someone without symptoms, but the 95% confidence interval for any symptoms overlaps the point estimate for no symptoms, and the two predictions are not statistically different. However, we see a much lower probability of going out for someone with other flu-like symptoms, relative to those with no COVID-19 symptoms. The bottom five bars in Figure 2 plot predicted probabilities for Model 2, where we estimate the effect of the three symptoms, fever, dry cough, or shortness of breath, separately. The results resemble those from Model 1. Relative to having no symptoms, having these three symptoms do not have a significant effect on the probability of going out, but having other flu-like symptoms continues to lower the probability of going out significantly.

**Figure 2.**
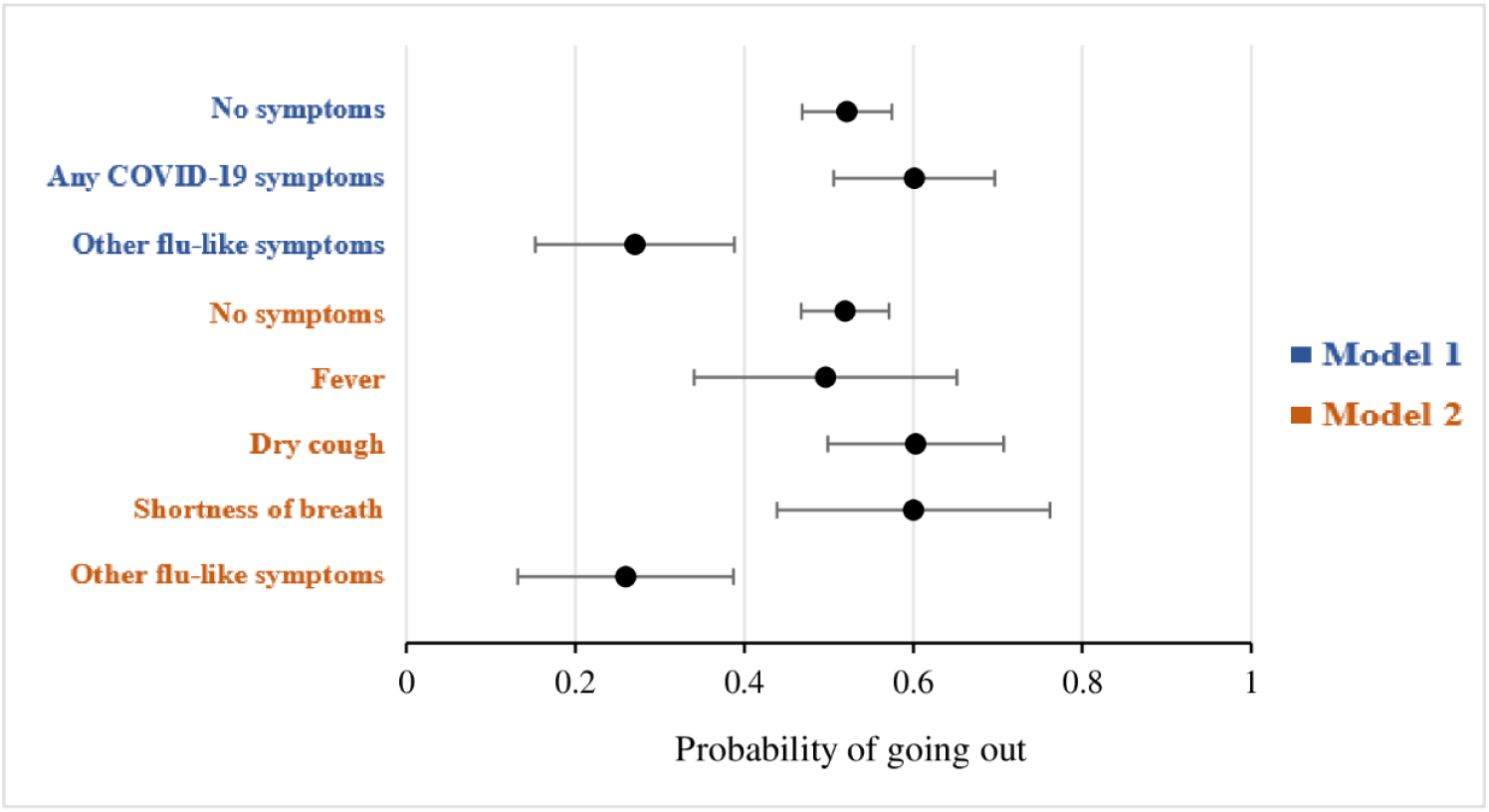
Predicted probabilities of going out yesterday by COVID-19 symptoms. Notes: (A) Yesterday refers to the day before completing the survey; (B) No symptoms refer to neither COVID-19 symptoms nor other flu-like symptoms; (C) Predicted probabilities and 95 percent confidence intervals were calculated at mean values of other covariates. (D) Probabilities are based on coefficients reported in Model 1 and Model 2 of Table 2.

Figure 3 plots the predicted probability of leaving the home yesterday, with 95% confidence intervals, by age group. These predicted probabilities are based on the coefficients reported in Model 1 of Table 2. Again, the predictions are made at the mean values of the other covariates and hence show the predicted probabilities by age, keeping other factors constant. We do not see any clear gradient in the predicted probabilities of going out of the home with age.

**Figure 3.**
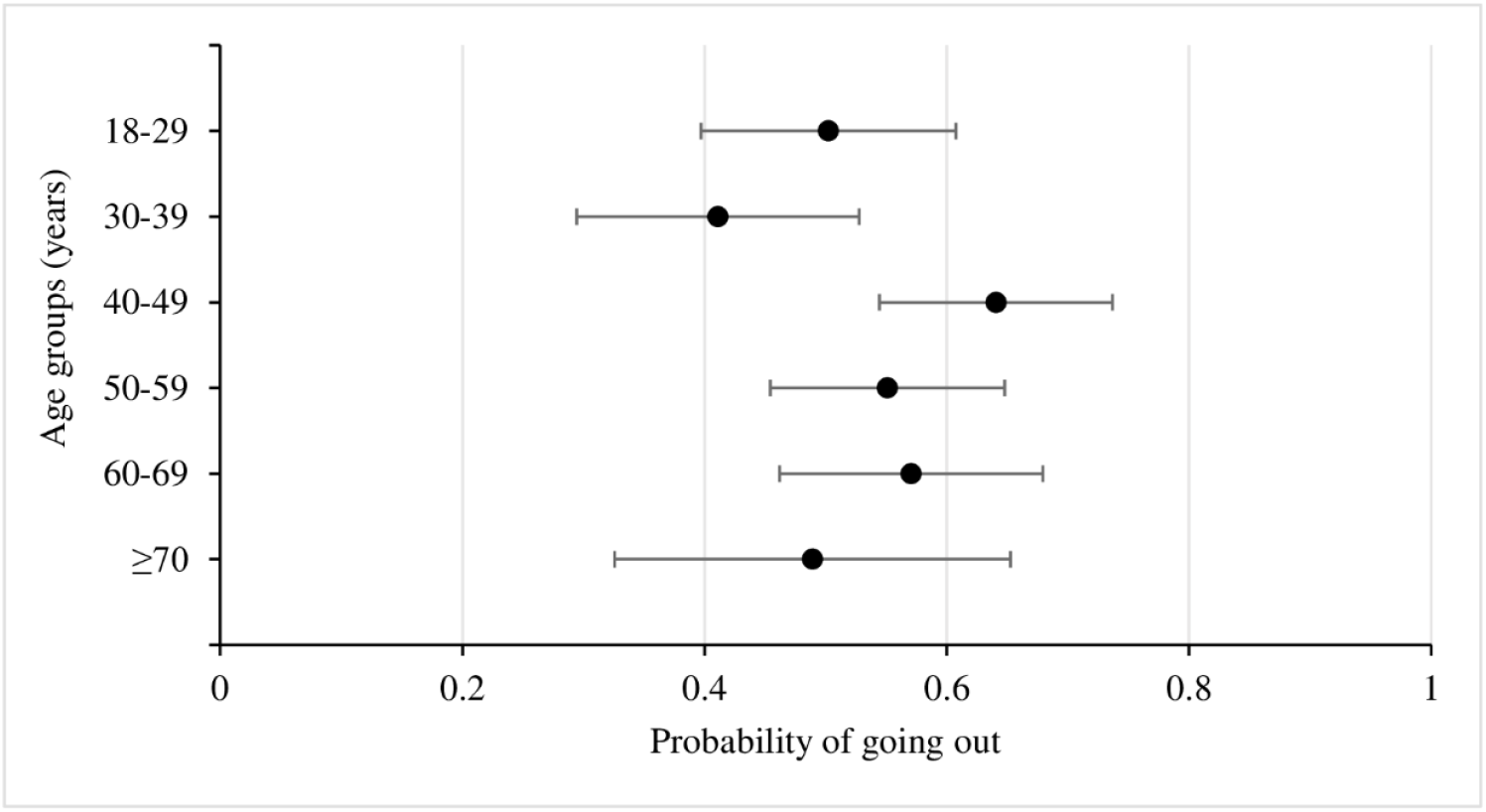
Predicted probabilities of going out yesterday by age group. Notes: (A) Yesterday refers to the day before completing the survey; (B) Predicted probabilities and 95 percent confidence intervals were calculated at mean values of other covariates. (C) Probabilities are based on coefficients reported in Model 1 of Table 2.

We predict the number of non-household members people come into close contact with (within 6 feet) using a Poisson count regression, and we report the results in Models 3 and 4 of Table 2. We again use sample weights. We use the same explanatory variables as in our models for leaving the home. The coefficient estimates in these models are reported as incidence rate ratios, and 95% confidence intervals are constructed using heteroskedastic robust standard errors. The results in Model 3 of Table 2 suggest that having any common symptom of COVID-19 is associated with a higher expected number of close contacts, but the effect does not have a statistically significant difference from a ratio of 1. However, we again find that other flu-like symptoms are significantly associated with more social distancing and having fewer close contacts, with an incidence rate ratio of 0.58 (95% CI: 0.38–0.88). We also find a strong age gradient; older people have fewer predicted close contacts that younger people. The oldest age group, those over 70 years of age, have an incidence rate ratio of 0.44 (95% CI: 0.34–0.81) relative to the youngest group (aged 18–29). We again find that those respondents who worked for pay in February 2020 tended to report less social distancing and a higher expected number of close contacts. We also find evidence of a gradient in income, with those with incomes more than $20,000 a year having more close contacts than those with lower incomes.

In Model 4, we again estimate the determinants the number of close contacts, but allow the three COVID-19 symptoms to have separate effects. While we find no significant effect of fever on the number of close contacts, we find that a dry cough raises the expected number of close contacts, with an incidence rate ratio of 1.95 (95% CI: 1.35– 2.81), while shortness of breath lowers the expected number of close contacts, with an incidence rate ratio of 0.48 (95% CI: 0.30–0.77). However, cough is a much more common symptom in the population (see Figure 1), which explains why any reporting of any symptom of COVID-19 tends to raise the expected number of contacts in Model 3. Results for other covariates are very similar to the results in Model 3.

Figure 4 shows the predicted number of close contacts, and their respective 95 percent confidence intervals, by COVID-19 symptoms in the last two weeks based on the estimates reported in Models 3 and 4 of Table 2. These predicted numbers of close contacts are calculated at the means of the other covariates. The predictions of Model 3 for the number of close contacts resemble the predictions of Model 1 for the probability of going out. Having any symptom of COVID-19 slightly raises the expected number of close contacts relative to those with no symptoms, but the confidence interval overlaps the estimate for those with no symptoms, and the difference is not statistically significant at the 5 percent level. However, other flu-like symptoms significantly lowers the expected number of close contacts. The predictions from Model 4 highlight the higher predicted number of contacts for those with a cough, though the confidence interval is quite wide, with a lower expected number of contacts with those reporting shortness of breath.

**Figure 4.**
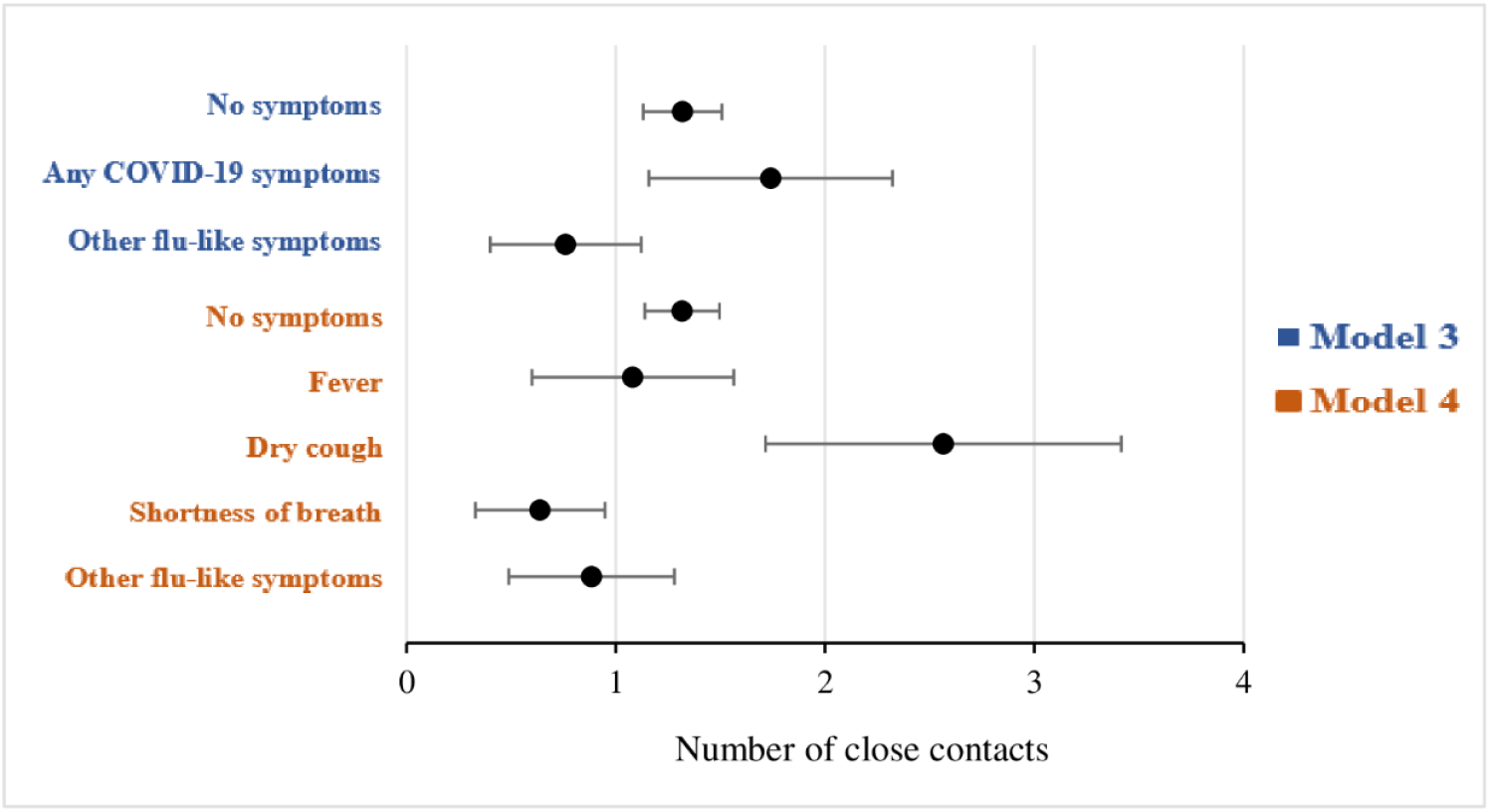
Predicted number of close contacts yesterday by COVID-19 symptoms. Notes: (A) Close contact means coming within six feet of a non-household member; (B) Yesterday refers to the day before completing the survey; (C) No symptoms refers to neither COVID-19 symptoms nor other flu-like symptoms. (D) Predicted number of contacts and 95 percent confidence intervals were calculated at mean values of other covariates. (E) Predicted numbers of close contacts are based on Model 3 and Model 4 of Table 2.

There is a clear age gradient in the number of close contacts, as shown in Figure 5, which is based on the results presented in Model 3 of Table 2. Those aged 18-29 are expected to come into contact with 2.3 non-household members per day, while those aged above 50 are predicted to come into close contact with less than half this number.

**Figure 5.**
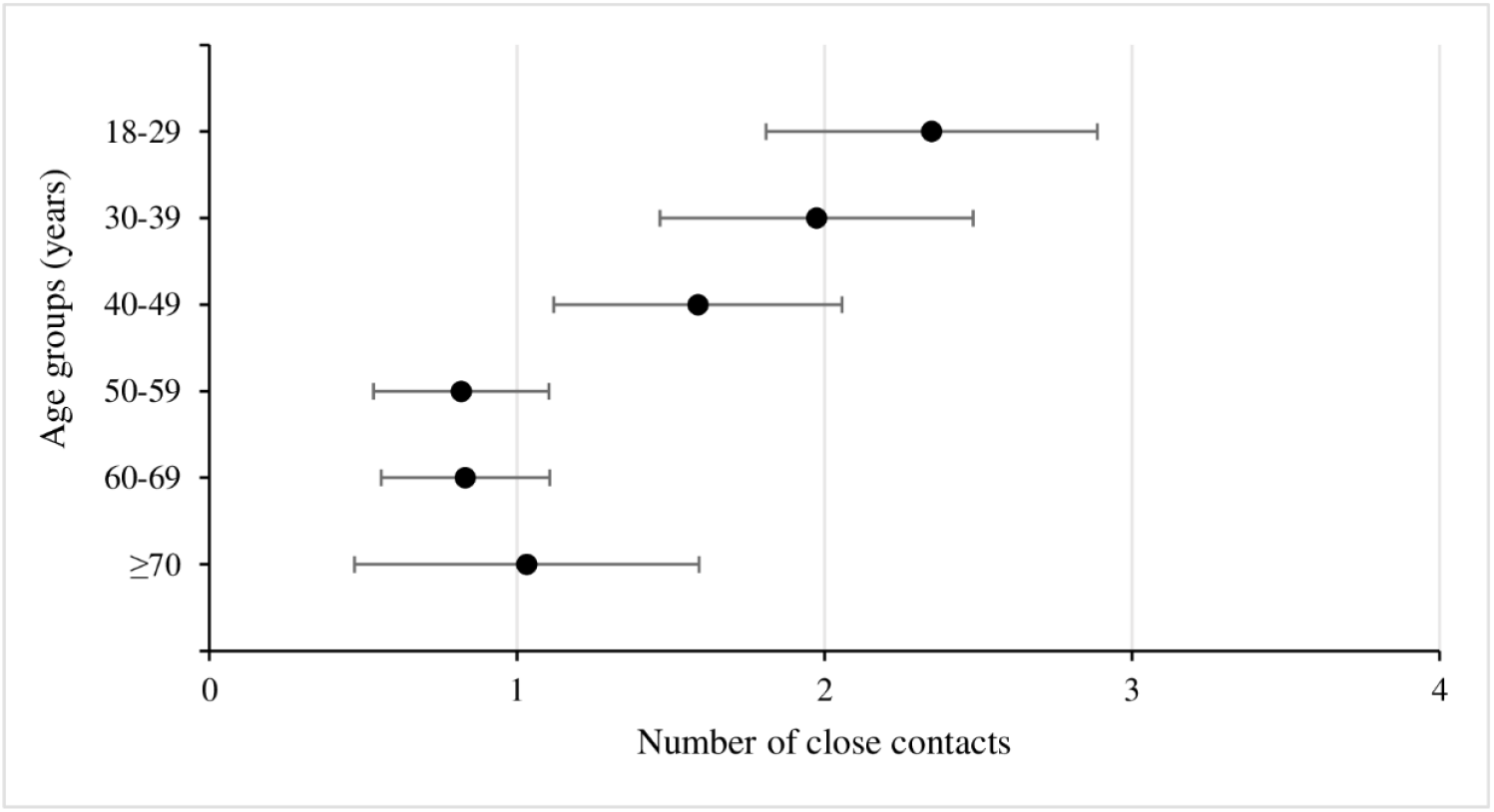
Predicted number of close contacts yesterday by age group. Notes: (A) Close contact means coming within six feet of a non-household member; (B) Yesterday refers to the day before completing the survey; (C) Predicted number of contacts and 95 percent confidence intervals were calculated at mean values of other covariates. (D) Predicted numbers of close contacts are based on Model 3 of Table 2.

## Discussion

We find a clear picture of how social distancing varies with age. Older people seem just as likely as younger people to go out of their homes. However, the expected number of close contacts with non-household members falls rapidly with age. Holding other factors constant, people over 50 years of age have less than half the expected number of close contacts than people age 18–29. This age gradient is in line with the WHO and CDC recommendations that older people should take special precautions, because they are more likely to develop a severe illness if they contract COVID-19. However, older adults in the United States tend to have smaller social networks compared with younger adults [24], which could also contribute to the phenomenon we are observing here.

Our findings with respect to COVID-19 symptoms are less clear. Using any common symptom of COVID-19 (fever, dry cough, or shortness of breath) in the last two weeks as the measure, we find no evidence of increased social distancing. However, when we disaggregate our exposure by COVID-19 symptoms, we do find that those adults who have had a cough have an increased expected number of close contacts, while those adults with shortness of breath have significantly fewer contacts, compared with adults without symptoms. We find consistently that those adults who had other flu-like symptoms, which are not routinely thought by WHO and CDC to be symptoms of COVID-19, are going out less, and are having fewer close contacts than others.

A possible explanation of these findings is that coughs are very common in the United States and have multiple causes [25]. While having a cough in the last two weeks has a high prevalence in our sample (see Figure 1), most of these cases are quite likely not due to COVID-19, but to other conditions. While cough is a common symptom of COVID-19, the probability of having COVID-19 given that someone has a cough may be very low. People may understand this intuitively and therefore may not be social distancing when they have a cough. Shortness of breath, and other flu-like symptoms, are comparatively rare, and people may be using these as stronger indicators that they may have COVID-19, and hence undertake greater social distancing. The lack of effect for fever, however, is surprising in this interpretation.

An alternative explanation of our results is that those adults who have a higher risk of severe illness are undertaking more intense social distance in their own interests, while those adults with COVID-19 symptoms are not, because any benefits accrue to others rather than to themselves. However, our evidence suggests that people with some symptoms, particularly shortness of breath and other flu-like symptoms, are undertaking more intense social distancing. Overall, the issue seems to be one of mismatch between the symptoms that people are using to inform their social distancing behavior and the symptoms of COVID-19 that are listed by the WHO and CDC. This mismatch could be a pure messaging issue, but the symptoms that are listed by the CDC and WHO could also lack sensitivity and specificity, and may not actually by thought by the public to be reliable guides to having COVID-19.

Our study has several limitations. One limitation is our measures of symptoms. Some of the symptoms of we measure may be chronic, and people may know that these symptoms are not related to COVID-19, and so may not be social distancing. Additionally, two weeks may be too long of a time frame; people may reduce their social distancing behavior only for a short while after symptoms end. A new study with greater temporal resolution of the timing of symptoms, which could control for chronic versus recent onset of symptoms, would be useful.

The selectivity of our sample is also an issue. A survey with greater balance of respondent characteristics would be desirable so that we do not have such extreme weights on some subgroups. Ideally, the sample would be a random sample of the population so that representativeness would be likely without the need to weight.

Our results may also be confounded by the fact that people who go out a lot are more likely to get symptoms of COVID-19, and this may lead to the positive association between symptoms and going out that we observe. We measure the symptoms over the last two weeks and going out only the day before, so this temporal ordering suggests that the causality runs from symptoms to going out, rather than vice versa. However, the possibility exists of unobserved individual characteristics that are not controlled for in our analysis that affect both going out and getting COVID-19 symptoms. This unobserved heterogeneity could be addressed through a panel data study using individual fixed effects, but we cannot make this adjustment using our cross-sectional data.

Our short survey fails to collect some information which could prove valuable. It would be desirable for a new survey to collect information on several other issues. This could include information on underlying health conditions that might put people at greater risk of severe illness; we focus only on the older population as a high-risk group. Additional demographic information, such as race and ethnicity, would be useful to see if these groups are at higher risk of infection. It would also be useful to collect data on other recommended prevention measures, such as handwashing and wearing a face covering in public, as additional outcomes.

Despite the limitations of the study design, it has enabled us to produce timely data that are relevant to a major public health emergency. We find that older people are undertaking greater social distancing, which is in line with public health recommendations. Our results are much less clear for enhanced social distancing by people who may have COVID-19 to prevent infections of others.

Our findings lead us to recommend two policy changes. The first recommendation is that in addition to listing the common symptoms of COVID-19, public health authorities should give clear guidance on what symptoms signal a strong likelihood of having COVID-19, and should lead to more intense social distancing, which may be quite different [19]. Ideally, a simple algorithm would calculate the probability of infection based on easily observable symptoms, with high specificity and sensitivity [19, 26]. The second recommendation is that messaging should be clear regarding how people should respond if they have symptoms and are likely to have COVID-19.

## Data Availability

Data collection of this study is still ongoing. Once complete and cleaned a data set for replication of results will be made available

## Acknowledgments

The authors would like to thank everyone who participated in the design, review, and response of this survey. The authors would also like to thank Kevin Gallagher, Adil Najam, and the support staff at the Program on the Global Demography of Aging at Harvard University and at the Global Development Policy Center at Boston University for their support on the development of the platforms for the web survey and on the dissemination and outreach to participants.

## Author Contributions

All authors participated equally in the conception, analysis, design, and writing of the article. All authors have read and approved the final manuscript and are aware that the manuscript is being submitted to the journal.

